# Anatomical Approach from the Coronary Sinus of Valsalva for Para-Hisian Premature Ventricular Contractions: Safety and Outcomes

**DOI:** 10.64898/2026.01.12.26343961

**Authors:** Masafumi Sugawara, Jakub Sroubek, Justin Lee, Bryan Baranowski, Mandeep Bhargava, Thomas Callahan, Mina Chung, Thomas Dresing, Ayman Hussein, Mohamed Kanj, Arshneel Kochar, Robert Koeth, David Martin, Walid Saliba, Tyler Taigen, Niraj Varma, Oussama Wazni, Pasquale Santangeli, Koji Higuchi

## Abstract

**Background:** While various reports described the characteristics and catheter ablation strategies for para-Hisian premature ventricular contraction (PVC), the patient population were limited in previous literatures and optimal approach remain unknown. This study sought to comprehensively assess the safety and outcomes of catheter ablation for para-Hisian PVC, with a particular focus on the safety and efficacy of ablation from aortic coronary cusps.

**Methods:** A total of 122 para-Hisian PVC cases were retrospectively included in this study. The acute and clinical outcomes of ablation, especially the success rate, the rate of conduction system injury, and the ablation site (right side, left side His bundle (HB) region, and aortic coronary cusp), were analyzed.

**Results:** Acute procedural success was achieved in 89 patients (73%). However, long-term clinical success (≥80% reduction of PVC burden) was achieved only in 52% of acute-success cases. Cusp ablation was attempted in 35 patients (31%). Cusp ablation was performed significantly more often in clinical-success cases than clinical-failed cases (38% vs.7% *P* = 0.005), although ablation from multiple locations was still required. In 28 of 35 cases (80%) who underwent cusp ablation, cusp ablation affected PVC by eliminating, transiently suppressing, or changing morphology. Predictors of successful cusp ablation included earlier activation at the cusp and close anatomical proximity to the earliest site at the HB region. Conduction system injury occurred in 31 cases of the entire cohort, mainly during the HB-region ablation, with only one event during cusp ablation.

**Conclusion:** In this large cohort of para-Hisian PVC ablation, an anatomical approach from the adjacent coronary cusp ablation can be considered as an effective and safe strategy. Early local activation and a close anatomical proximity between the earliest para-Hisian site and the coronary cusp region are predictors of ablation success with this approach.

**What is known:**

- Catheter ablation of para-Hisian premature ventricular contraction (PVC) is challenging due to the proximity to the His bundle (HB) and/or potentially intramural sources.
- Catheter mapping and ablation from multiple sites, including aortic coronary cusps, have been reported as potential alternative approaches, however systematic analyses of the ablation outcome and safety for para-Hisian PVC remain scarce.

**What the study adds:**

- Clinical-success rate of radiofrequency (RF) ablation for para-Hisian PVC was still suboptimal (52%). However, the cases undergoing cusp ablation were associated with a better success rate (85%) than those with direct ablation to the HB region alone (42%), indicating this approach may improve the outcome.
- The anatomical features, including PVC earliest activation site above the HB or within short distance from the coronary cusp, may predict successful PVC elimination from the coronary cusp.
- Conduction system injury, including minor forms, occurred in approximately 30% cases during the RF ablation near the HB region, while only one AH prolongation observed during cusp ablation, indicating superior safety of cusp ablation.

## Introduction

Premature ventricular contractions (PVCs) originating from the His bundle (HB) region, known as para-Hisian PVCs, account for approximately 3–9% of idiopathic ventricular arrhythmias and may also occur in patients with structural heart disease. ^1–3^ Radiofrequency (RF) ablation at the earliest activation site (EAS) near the HB region carries a significant risk of damaging the cardiac conduction system, which may lead to a suboptimal procedural success rate. To improve both the efficacy and safety of ablation for para-Hisian PVCs, various alternative strategies, such as ablation from the aortic coronary cusps or an anatomical approach from the contralateral chamber, have been proposed. ^4–9^

Despite being relatively common, ablation of para-Hisian PVCs is often deferred due to the related risks, leading to a limited number of reported cases. Consequently, comprehensive evaluations of procedural success and conduction system injury incidence remain scarce, and there remains a need for further investigation to determine the optimal ablation strategy for para-Hisian PVC.

The aim of this study is to evaluate the acute success rate, long-term recurrence rate and incidence of conduction system injuries through comprehensive analyses of a large consecutive series of para-Hisian PVC cases undergoing catheter mapping and ablation, with a particular focus on the impact of anatomical ablation from the adjacent aortic coronary cusp.

## Methods

### Study design and patient population

This is a single-center retrospective cohort study reviewing consecutive 1,620 patients who underwent catheter ablation for PVC between January 2015 and August 2024 at the Cleveland Clinic, Cleveland, OH. Para-Hisian PVC cases were identified in these cases based on the surface electrocardiogram (ECG) criteria and electro-anatomical mapping (EAM) findings described below. Cases with multiple clinical PVCs or ventricular-pacing dependence were excluded. Ablation outcomes and incidence of conduction system injury were analyzed based on electrocardiographic findings and electrophysiological data obtained during the procedure. The study protocol was approved by the Institutional Review Board and conducted in accordance with the Helsinki Declaration as revised in 2013.

### Definition of para-Hisian PVC

The para-Hisian PVC was identified based on both of (1) ECG features (**Figure 1A**) ^1,3,10,11^ and (2) EAM with the earliest PVC activation located within 10 mm from the closest HB potential recording site (**Figures 1B and 1C**). ^3–6,12–15^

**Figure 1.**
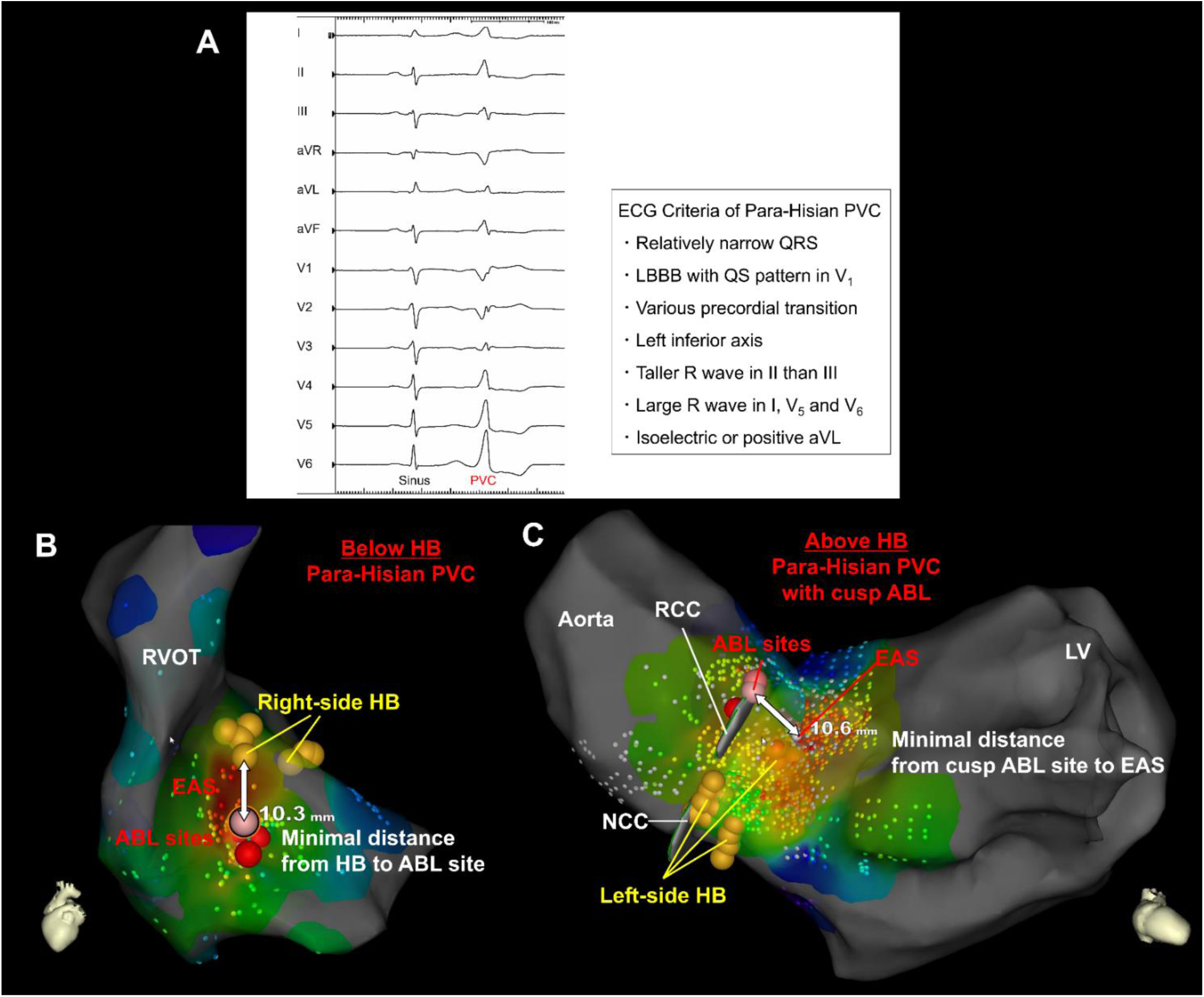
Representative ECG and EAM of para-Hisian PVC. **(A)** Representative electrocardiogram (ECG) showing a para-Hisian premature ventricular contraction (PVC) and its diagnostic criteria. **(B)** Representative electroanatomical map (EAM) of a right-sided below-His para-Hisian PVC. The earliest activation site (EAS) was localized below the His bundle (HB) (yellow tags). Ablation (red tags) was delivered inferior to the EAS, with a minimal distance of 10.3 mm from the HB. **(C)** Representative EAM of a left-sided above-His para-Hisian PVC. The EAS was localized above the HB, and ablation was delivered in the right coronary cusp (RCC) with a minimal distance of 10.6 mm from the EAS. LBBB indicates left bundle branch block; LV, left ventricule; NCC, non-coronary cusp; and RVOT, right ventriclar outflow tract.

### Mapping and ablation procedure for para-Hisian PVC

All antiarrhythmic drugs were discontinued before catheter mapping and ablation. Procedures were performed under conscious sedation using a three-dimensional (3D) mapping system (CARTO 3; Johnson&Johnson MedTech, Irvine, CA, or EnSite; Abbott, Chicago, IL) with compatible multi-electrode and irrigated contact-force RF ablation catheters. When clinical PVCs were infrequent, programmed stimulation or isoproterenol infusion was used to induce PVCs. Intravenous heparin was administered to maintain an activated clotting time >350 ms before mapping in the aortic cusps or left ventricle (LV). Activation mapping was first performed in the right ventricle (RV) and LV near the HB. The local activation time was annotated by the wavefront algorithm when CARTO 3 system was used, and the first deflection of the ventricular potential on bipolar electrogram (EGM) when EnSite system was used. ^16^ Discrete near-field HB potentials were tagged on the EAM. Mapping was further extended to aortic coronary cusps as needed based on the operator’s discretion. RF energy was delivered at 10–50 W with a target contact force of 10–20 g, with careful monitoring for potential conduction system injury. During ablation within the coronary cusps, catheter position was continuously assessed using intracardiac echocardiography to confirm a safe distance from the coronary artery ostia. Following ablation, PVC burden was monitored for approximately 30 minutes to evaluate the acute procedural outcome. Mapping and ablation were repeated at alternative sites until acute success was achieved or the procedure was terminated at the operator’s discretion.

### Definitions of clinical outcomes and incidence of conduction system injury

Acute success was defined as suppression of the clinical PVC during the procedural observation period, as determined by the operator. This was evaluated in all cases in which ablation was attempted (acute-outcome cohort). Clinical success was defined as a reduction in PVC burden of ≥80%, assessed using a 24- or 48-hour Holter ECG or a 2-week heart monitor during a follow-up period, as described in previous literature. ^2,17,18^ This outcome was evaluated only for cases with acute procedural success and available follow-up data (clinical-outcome cohort).

The incidence of new-onset atrioventricular (AV) conduction injury was categorized into four types: (1) AH/HV prolongation or first-degree AV block (PR interval > 200 msec), (2) right bundle branch block (RBBB), (3) left bundle branch block (LBBB) or left anterior fascicular block (LAFB) (4) Advanced AV block (AVB), including 2:1 AVB or complete AVB. Conduction injury that resolved to baseline during hospitalization was classified as “transient”, whereas injury persisting until discharge was classified as “permanent”.

### Retrospective analysis of electrophysiological characteristics

The PVC activation time (ms), defined as the interval from the local ventricular bipolar EGM to the onset of the surface QRS complex (V–QRS), was manually measured retrospectively using each 3D mapping system. Local bipolar EGM annotation was performed as described above. The EAS for each chamber was identified using activation maps generated from all mapped chambers.

Cases were classified as above HB or below HB, according to the vertical anatomical location of the EAS on the 3D mapping. Above-HB was defined as an EAS located superior to the HB and in close proximity to the coronary cusps, whereas below-HB was defined as an EAS located inferior to the HB and remote from the coronary cusps (**Figures 1B and 1C**). ^15^

Mapping and ablation sites were categorized into three locations: (1) right-side HB region, (2) left-side HB region, and (3) aortic coronary cusps. The location and sequence of the mapped and ablated sites were evaluated for each case. For each patient, the total number of RF applications, maximum RF power (W), and maximum ablation duration (sec) were analyzed in relation to the procedural success.

### Detailed analysis of aortic cusp mapping and ablation

Cases that underwent mapping and ablation in the aortic coronary cusps were separately analyzed. The effects of cusp ablation were further evaluated and categorized into four groups: (1) PVC termination, (2) transient PVC suppression, (3) change in PVC morphology, and (4) minimal impact. The effects of cusp ablation were determined by reviewing surface ECGs recorded during each RF application at the aortic cusp using the recording system (CardioLab; GE HealthCare, Chicago, IL). The V–QRS interval at the EAS within the aortic cusp was manually measured as described above. In cases undergoing cusp ablation, the minimal distance from the cusp ablation site to the EAS at the HB region was measured on the 3D mapping system (**Figure 1C**). These parameters (V–QRS interval and anatomical distance) were then compared according to the observed effects of cusp ablation.

### Detailed analysis for incidence of conduction system injury

Cases in which conduction system injury occurred during the procedure were separately analyzed. The RF application site responsible for the conduction disturbance was classified as one of the following: (1) right-sided HB region, (2) left-sided HB region, or (3) aortic coronary cusp. The minimal distance from the ablation site to the closest near-field HB potential recording site was measured on the 3D mapping system (**Figure 1B**). In addition, the presence or absence of far-field HB potentials recorded at the ablation site was evaluated and compared between cases with and without conduction system injury.

### Statistical analysis

Continuous variables were expressed as mean ± standard deviation (SD) or median with interquartile range (IQR), while categorical variables were presented as numbers with percentage (%). Two-group comparisons were conducted using an unpaired Student’s t-test or Mann-Whitney U test for continuous variables, and a chi-square test or Fisher’s exact test for categorical variables, as appropriate. A *P*-value of <0.05 was considered statistically significant. All statistical analyses were performed using JMP software version 16.2.0 (SAS Institute, Cary, NC).

## Results

### Acute and long-term ablation outcome

A total of 122 cases of para-Hisian PVCs were identified and retrospectively analyzed. The procedure was aborted in 8 cases without any ablation attempts due to the proximity to the conduction system. Among the 114 cases in which ablation was performed (acute-outcome cohort), acute procedural success was achieved in 89 cases, while 25 cases resulted in acute failure (**Figure 2A**). Ablation was attempted from the coronary cusps in 35 patients (31%). Of the 56 acute-success cases with available follow-up data (clinical-outcome cohort, median follow-up duration: 65.5 days [IQR 21-450]), 29 cases achieved clinical success, defined as a ≥80% reduction in PVC burden, whereas 27 cases showed a recurrence of PVC (**Figure 2B**). Baseline characteristics of the entire cohort and each outcome subgroup are summarized in **Table 1**. Detailed mapping and ablation locations, as well as procedural sequences for each case, are illustrated as flowcharts in **Supplemental Figures 1–4**.

**Figure 2.**
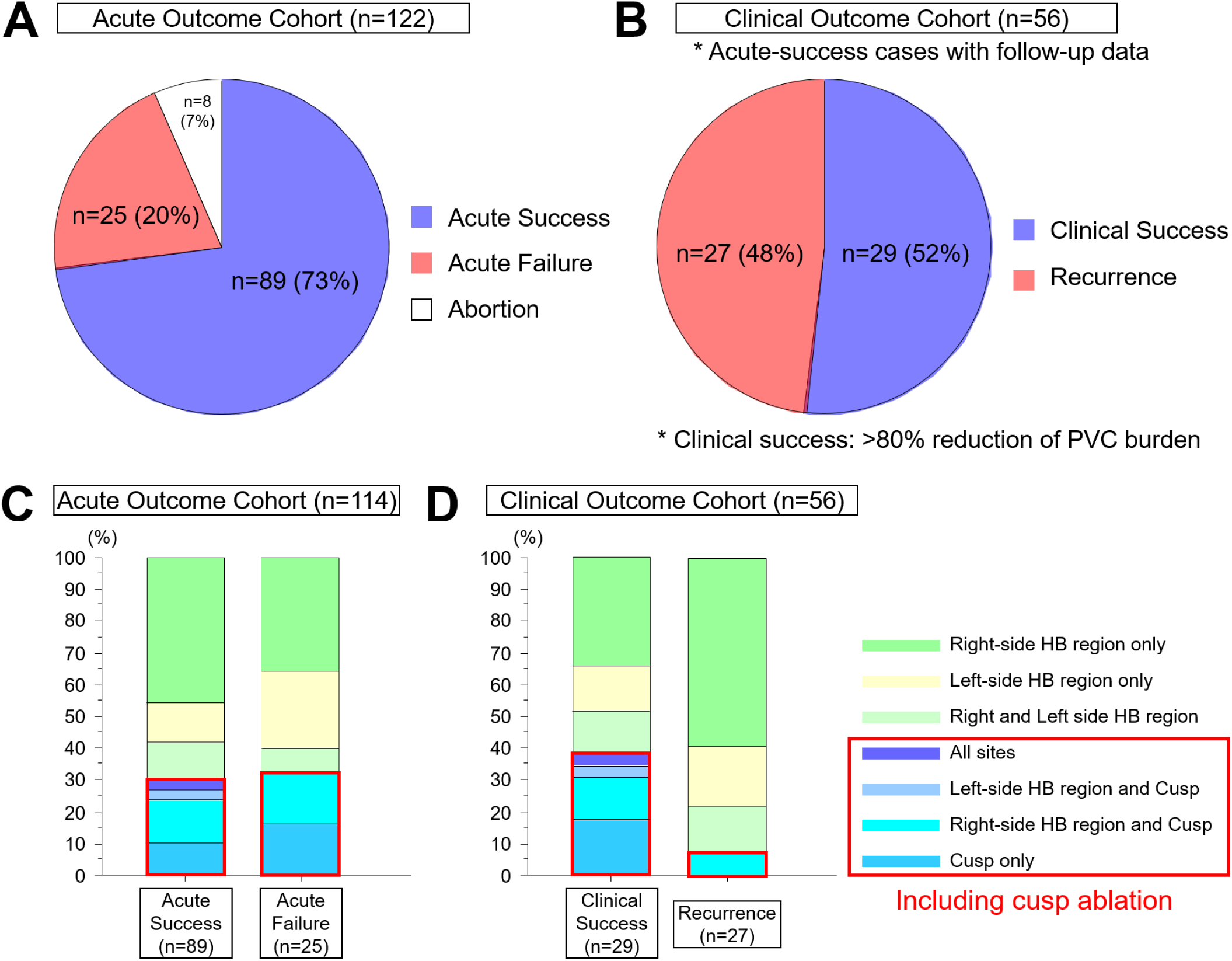
Acute and long-term outcome after ablation of para-Hisian PVC. **(A)** Success rate in the acute-outcome cohort. **(B)** Success rate in the clinical-outcome cohort. **(C)** Distribution of ablation sites in the acute-outcome cohort. **(D)** Distribution of ablation sites in the clinical-outcome cohort. Red boxes indicates patiens who had cusp ablation. Cusp ablation was performed in 38% of cases with clinical success (n = 11/29) compared with 7% of cases with premature ventricular contraction (PVC) recurrence (n = 2/27).

**Table 1.** Patients and procedural characteristics.

|  | Entire Cohort (n=122) | Acute-outcome cohort (n=114) |  |  | Clinical-outcome cohort (n=56) |  |  |
| --- | --- | --- | --- | --- | --- | --- | --- |
|  |  | Acute success (n=89) | Acute failure (n=25) | P-value | No Recurrence (n=29) | Recurrence (n=27) | P-value |
| Patient characteristics |  |  |  |  |  |  |  |
| Age, years | 63.1±13.4 | 62.2±14.1 | 64.1±11.8 | NS | 62.2±14.4 | 66.4±9.5 | NS |
| Male gender, n | 76 (62) | 53 (60) | 17 (68) | NS | 17 (59) | 12 (44) | NS |
| Body mass index, kg/m2 | 29.4±4.9 | 29.1±5.0 | 29.5±5.1 | NS | 28.9±4.9 | 29.4±5.7 | NS |
| Left ventricular ejection fraction, % | 51.1±11.8 | 50.1±11.6 | 53.2±13.9 | NS | 52.3±9.3 | 48.5±12.9 | NS |
| Underlying heart disease, n | 37 (30) | 31 (35) | 6 (24) | NS | 7 (24) | 13 (48) | NS |
| History of prior PVC ablation, n | 35 (29) | 23 (26) | 11 (44) | NS | 8 (28) | 4 (15) | NS |
| PVC burden |  |  |  |  |  |  |  |
| Pre PVC burden, % | 23.5±12.5 | 24.7±12.8 | 20.3±11.8 | NS | 25.3±10.9 | 24.7±13.8 | NS |
| Post PVC burden, % | 9.3±10.4 | NA | NA | NA | 0.9±1.6 | 16.1±8.9 | <0.001 |
| ECG characteristics of PVC |  |  |  |  |  |  |  |
| QRS duration, msec | 133±19 | 134±18 | 128±19 | NS | 136±19 | 131±18 | NS |
| Maximum Deflection Index | 0.56±0.1 | 0.56±0.1 | 0.54±0.1 | NS | 0.56±0.1 | 0.54±0.1 | NS |
| Transition in precordial leads |  |  |  |  |  |  |  |
| V1-V2, n | 31 (25) | 17 (19) | 11 (44) | 0.017 | 6 (21) | 6 (22) | NS |
| V2-V3, n | 45 (37) | 33 (37) | 10 (40) | NS | 10 (34) | 8 (30) | NS |
| V3-V4, n | 28 (23) | 23 (26) | 4 (16) | NS | 7 (24) | 7 (26) | NS |
| V4-V5, n | 17 (14) | 15 (17) | 0 (0) | 0.039 | 5 (17) | 6 (22) | NS |
| V5-V6, n | 1 (1) | 1 (1) | 0 (0) | NS | 1 (3) | 0 (0) | NS |
| Earliest activation site at HB region |  |  |  |  |  |  |  |
| Right side, n | 88 (72) | 66 (74) | 15 (60) | NS | 19 (66) | 21 (78) | NS |
| Left side, n | 34 (28) | 23 (26) | 10 (40) | NS | 10 (34) | 6 (22) | NS |
| Above His bundle, n | 53 (43) | 41 (46) | 11 (44) | NS | 15 (52) | 7 (26) | 0.059 |
| Below His bundle, n | 69 (57) | 48 (54) | 14 (56) | NS | 14 (48) | 20 (74) | 0.059 |
| Earliest V-QRS at HB region, msec | -24.8±9.4 | -25.8±9.7 | -22.6±8.9 | NS | -26.0±8.6 | -24.6±11.1 | NS |
| Ablation site (s), n |  |  |  |  |  |  |  |
| Right-side HB region only | 50 (41) | 41 (46) | 9 (36) | NS | 10 (34) | 16 (59) | NS |
| Left-side HB region only | 17 (14) | 11 (12) | 6 (24) | NS | 4 (14) | 5 (19) | NS |
| Right and Left-side HB region | 12 (10) | 10 (11) | 2 (8) | NS | 4 (14) | 4 (15) | NS |
| Coronary cusp only | 13 (11) | 9 (10) | 4 (16) | NS | 5 (17) | 0 (0) | 0.052 |
| Right-side HB region + Cusp | 16 (13) | 12 (13) | 4 (16) | NS | 4 (14) | 2 (7) | NS |
| Left-side HB region + Cusp | 3 (2) | 3 (3) | 0 (0) | NS | 1 (3) | 0 (0) | NS |
| Right and Left-side HB region + Cusp | 3 (2) | 3 (3) | 0 (0) | NS | 1 (3) | 0 (0) | NS |
| Abortion without any ablation attempt | 8 (7) | NA | NA | NA | NA | NA | NA |
| Ablation characteristics |  |  |  |  |  |  |  |
| Total number of RF application | 11.7±7.7 | 12.1±7.8 | 10.2±7.5 | NS | 12.4±8.9 | 11.7±6.2 | NS |
| Maximum RF power, Watt | 38.0±6.7 | 38.6±5.9 | 36.1±9.2 | NS | 38.3±4.5 | 38.8±5.9 | NS |
| Maximum RF duration, sec | 109.6±52.5 | 115.0±52.6 | 88.7±47.8 | 0.067 | 106.8±44.6 | 109.3±56.9 | NS |
| Major Complications |  |  |  |  |  |  |  |
| Cardiac tamponade, n | 0 (0) | 0 (0) | 0 (0) | NS | 0 (0) | 0 (0) | NS |
| Groin hematoma, n | 1 (1) | 1 (1) | 0 (0) | NS | 0 (0) | 1 (4) | NS |
| Pseudoaneurysm, n | 2 (2) | 1 (1) | 1 (4) | NS | 0 (0) | 0 (0) | NS |
| Incidence of AV conduction injury |  |  |  |  |  |  |  |
| Transient AH prolongation, n | 1 (1) | 1 (1) | 0 (0) |  | 1 (3) | 0 (0) |  |
| Transient 1st degree AVB, n | 2 (2) | 0 (0) | 2 (8) |  | 0 (0) | 0 (0) |  |
| Permanent 1st degree AVB, n | 1 (1) | 1 (1) | 0 (0) |  | 0 (0) | 1 (4) |  |
| Transient RBBB, n | 3 (2) | 3 (3) | 0 (0) |  | 1 (3) | 1 (4) |  |
| Permanent RBBB, n | 9 (7) | 8 (9) | 1 (4) |  | 2 (7) | 4 (15) |  |
| Transient LBBB/LAFB, n | 5 (4) | 5 (6) | 0 (0) |  | 2 (7) | 2 (7) |  |
| Permanent LBBB/LAFB, n | 2 (2) | 0 (0) | 2 (8) |  | 0 (0) | 0 (0) |  |
| Transient advanced AVB, n | 5 (4) | 3 (3) | 2 (8) |  | 2 (7) | 1 (4) |  |
| Permanent advanced AVB, n | 3 (2) | 3 (3) | 0 (0) |  | 0 (0) | 1 (4) |  |
| Total, n | 31 (25) | 24 (27) | 7 (28) | NS | 8 (28) | 10 (37) | NS |

AVB = atrioventricular block; ECG = electrocardiogram; HB = His bundle; LAFB = left anterior fascicular block; LBBB = left bundle branch block; PVC = premature ventricular contraction; RBBB = right bundle branch block; RF = radiofrequency.

### Factors associated with ablation success

The anatomical locations where RF ablation was performed are summarized in **Figures 2C and 2D**, categorized by procedural and clinical outcomes. Cusp ablation was performed in 38% of cases with clinical success (11 of 29) compared with 7% of cases with PVC recurrence (2 of 27) (**Figure 2D**, *P* = 0.005). Ablation parameters, including the total number of RF applications, maximum RF power, and maximum ablation duration, did not differ significantly between outcome groups in either cohort (**Figures 3A–F**, **Table 1**). At follow-up (clinical outcome cohort), patients who underwent ablation including coronary cusps achieved a significantly higher rate of successful outcome compared with those without cusp ablation (11/13 [85%] vs. 18/43 [42%], *P* = 0.005) (**Figure 4B**).

**Figure 3.**
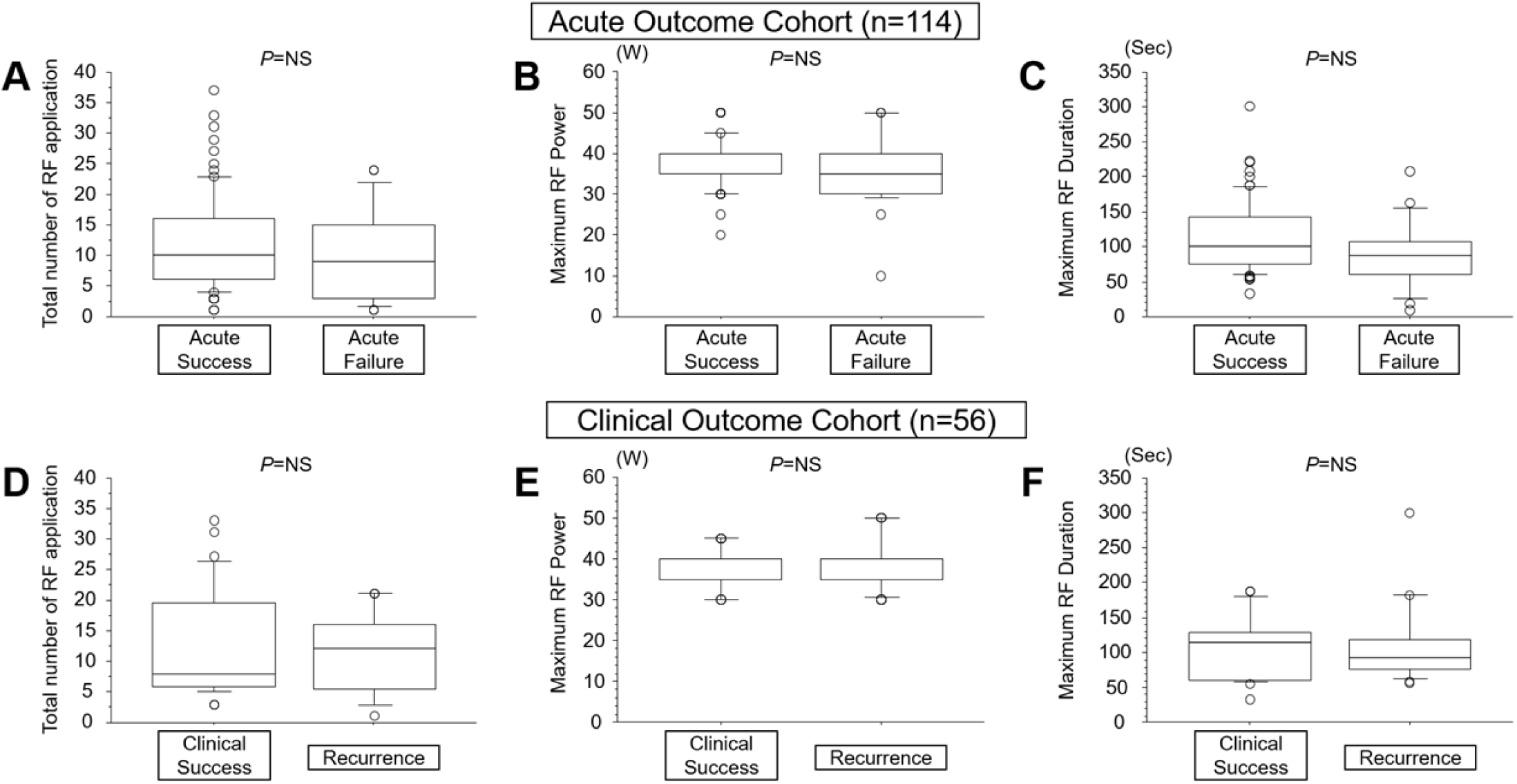
Comparison of the ablation parameters bewteen each outcome in each cohort. **(A)** Total number of radiofrequency (RF) applications in the acute-outcome cohort. **(B)** Maximum RF power in the acute-outcome cohort. **(C)** Maximum RF duration in the acute-outcome cohort. **(D)** Total number of RF applications in the clinical-outcome cohort. **(E)** Maximum RF power in the clinical-outcome cohort. **(F)** Maximum RF duration in the clinical-outcome cohort. No statistically significant differences were observed in any of the comparisons.

**Figure 4.**
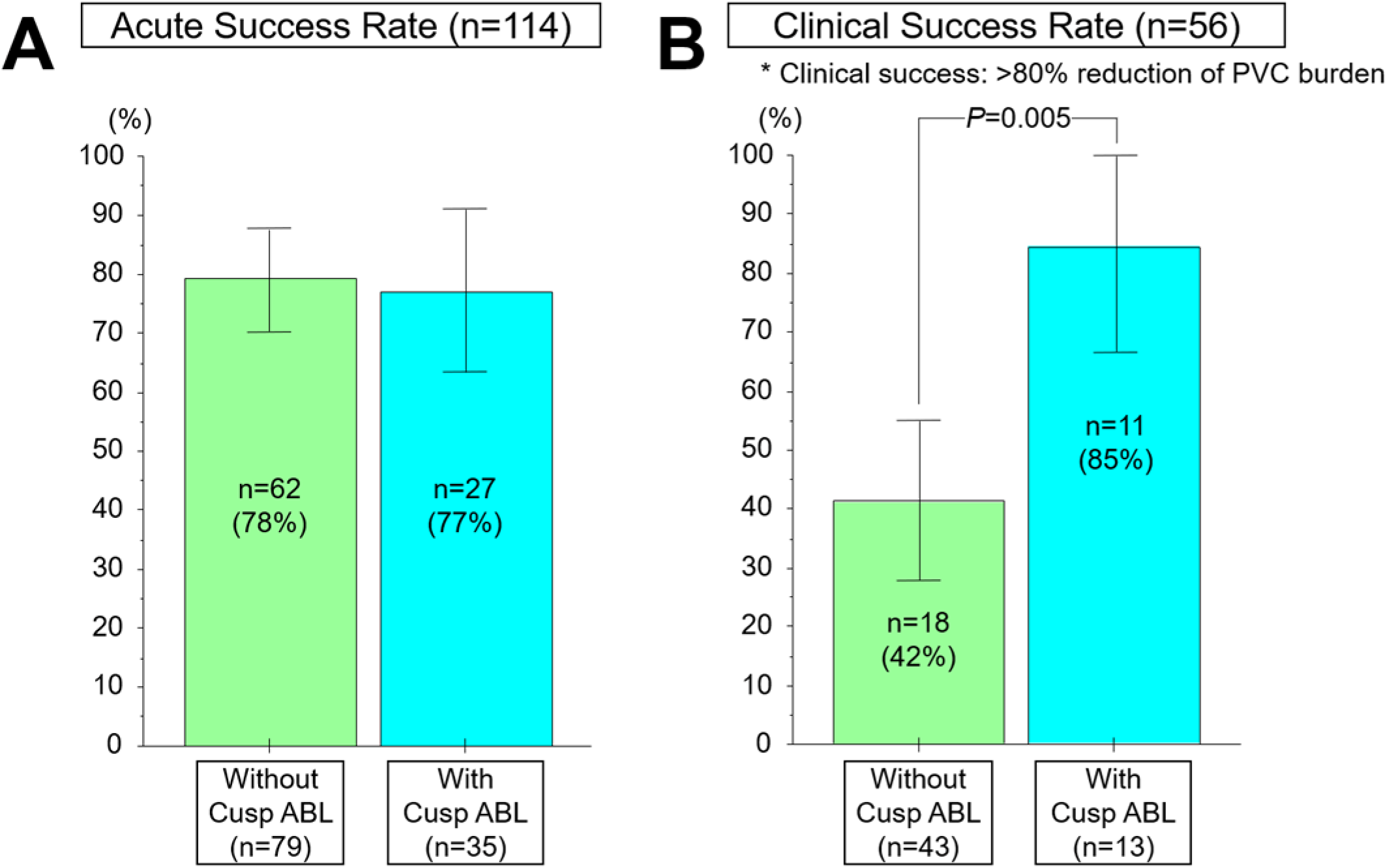
Comparison of the success rate between cases with and without cusp ablation. **(A)** Acute success rate. **(B)** Clinical success rate was significantly higher in the cases with cusp ablation (n = 11/13; 85%), compared with those without cusp ablation (n = 18/43; 42%) (P=0.005).

### Detailed analysis of cases with aortic cusp ablation

A subgroup analysis was performed in 51 cases in which catheter mapping was conducted within the aortic cusps (**Table 2**). In 16 of these cases, RF ablation was not attempted (activation time in the cusp was not early). Among the 35 cases undergoing cusp ablation, an effect on PVCs was observed in 80% (28 of 35) (**Figure 5A**). Cusp ablation resulted in complete PVC suppression in 17 cases. In 11 cases, ablation within the coronary cusps resulted in transient PVC suppression or a change in PVC morphology but failed to achieve complete elimination, necessitating additional ablation in the HB region. Cases with PVC termination by cusp ablation exhibited significantly earlier activation at the cusp compared with those without termination (V–QRS interval: −28.8 ± 10.2 ms vs. −14.8 ± 14.4 ms; *P* = 0.002) (**Figure 5B**). In all 17 successful cases, the EAS in the HB region was located above the HB. Furthermore, the minimal distance from the cusp ablation site to the EAS in the HB region was shorter in cases with successful PVC termination (9.1 ± 2.0 mm vs. 11.7 ± 2.9 mm; *P* = 0.004) (**Figure 5C**). **Figure 6** illustrates a representative para-Hisian PVC case in which cusp ablation modified PVC morphology. The EAS was located above the HB, and the right coronary cusp (RCC) demonstrated an activation time comparable to the right-sided HB region (−18 ms vs. −21 ms, respectively), suggesting an intramural origin (**Figure 6A**). RF ablation at the RCC, 10.9 mm from the EAS at right-sided HB region, transiently suppressed the PVC (**Figure 6B**), but was followed by recurrence with slightly altered morphology and reduced frequency. The increased S wave in the inferior leads indicated an inferior shift of the PVC exit (**Figure 6C**). Ultimately, RF ablation at the right-side HB EAS successfully eliminated the PVC (**Figure 6D**).

**Figure 5.**
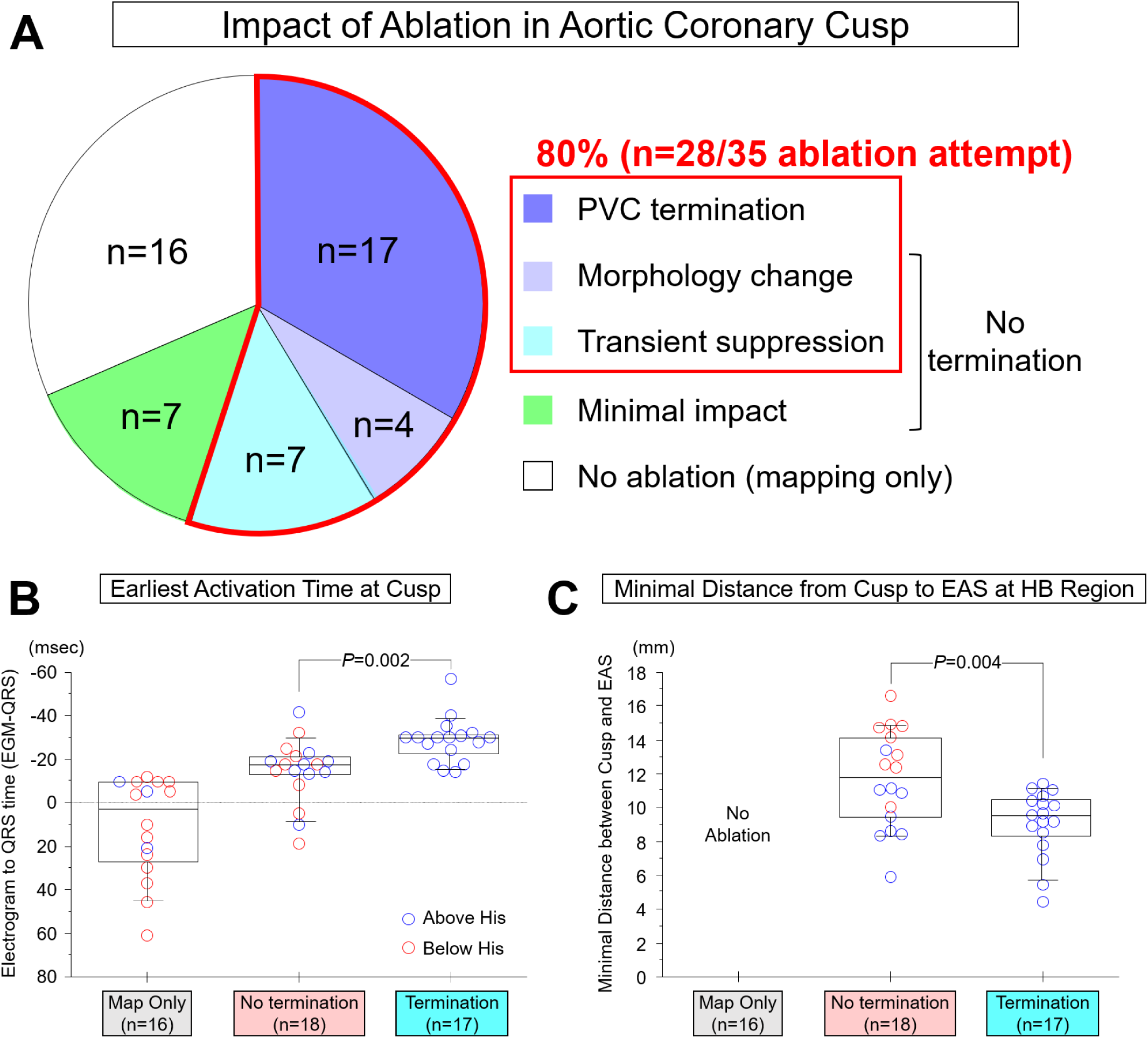
Impact of cusp ablation on the ablation outcome for para-Hisian PVC. **(A)** Distribution of outcomes following cusp ablation. Cusp ablation affected either the frequency or morphology of premature ventricular contractions (PVCs) in 80% of cases in which ablation was attempted (n = 28/35). **(B)** Comparison of PVC activation time between cases with and without successful cusp ablation. **(C)** Comparison of the distance from the cusp ablation site to the earliest activation site (EAS) in the His bundle (HB) region.

**Figure 6.**
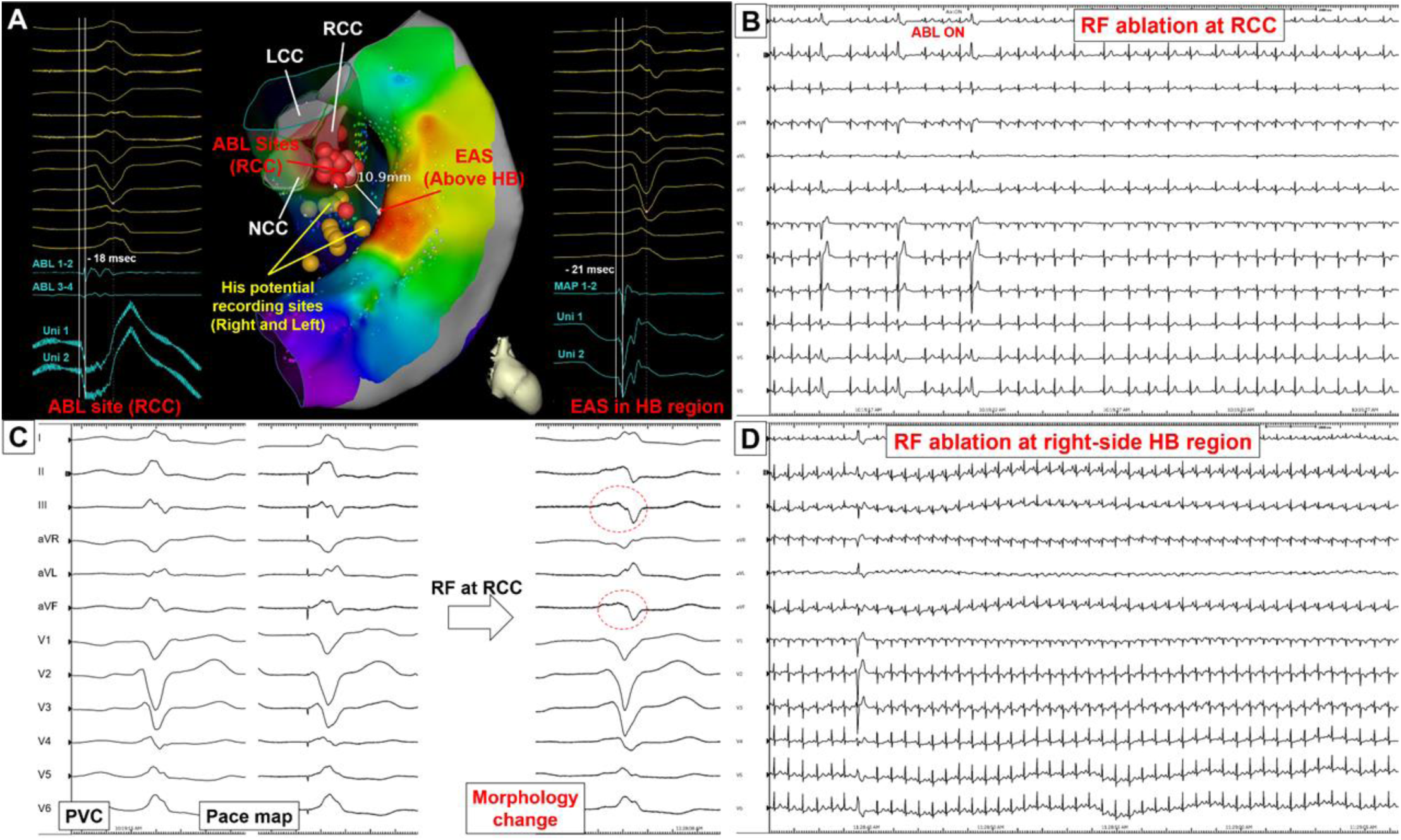
A representative para-Hisian PVC case in which cusp ablation influenced PVC frequency and morphology. See the main text for the details. LCC indicates left coronary cusp; RF, radiofrequency; and other abbreviations as in Figure 1.

**Table 2.**
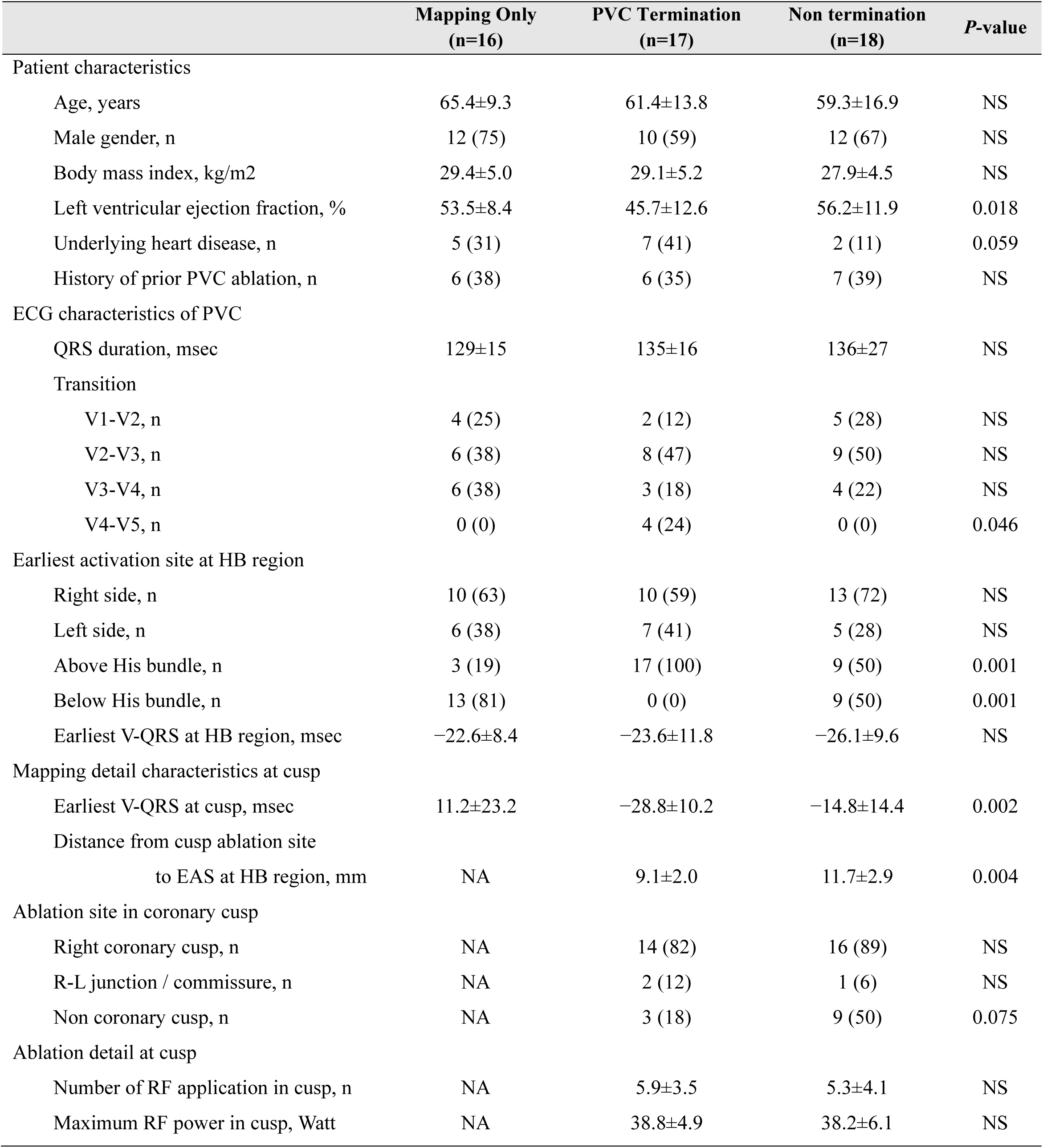
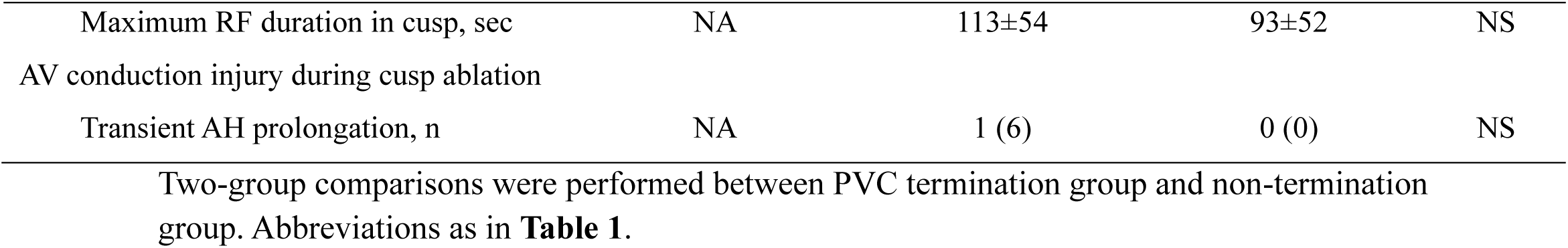
Patients and characteristics of cases underwent coronary cusp ablation.

### Incidence of conduction system injury

Among 114 cases undergoing ablation, a total of 31 periprocedural conduction system injuries were observed (**Figure 7A**, **Table 1**). Among 81 cases in which RF ablation was performed in the right-sided HB region (n = 50, right side only; n = 31, right side plus other site[s]), 19 conduction system injuries occurred (23%) during the right-sided HB-region ablation, including one case of permanent complete AVB. In 35 cases with RF ablation in the left-sided HB region (n = 17, left side only; n = 18, left side plus other site[s]), 11 conduction system injuries occurred (31%) during the left-sided HB-region ablation, also including one permanent complete AVB. In contrast, among 35 cases with RF ablation performed in the aortic cusps, only one conduction system injury occurred (3%) during cusp ablation, and no permanent injuries were observed.

**Figure 7.**
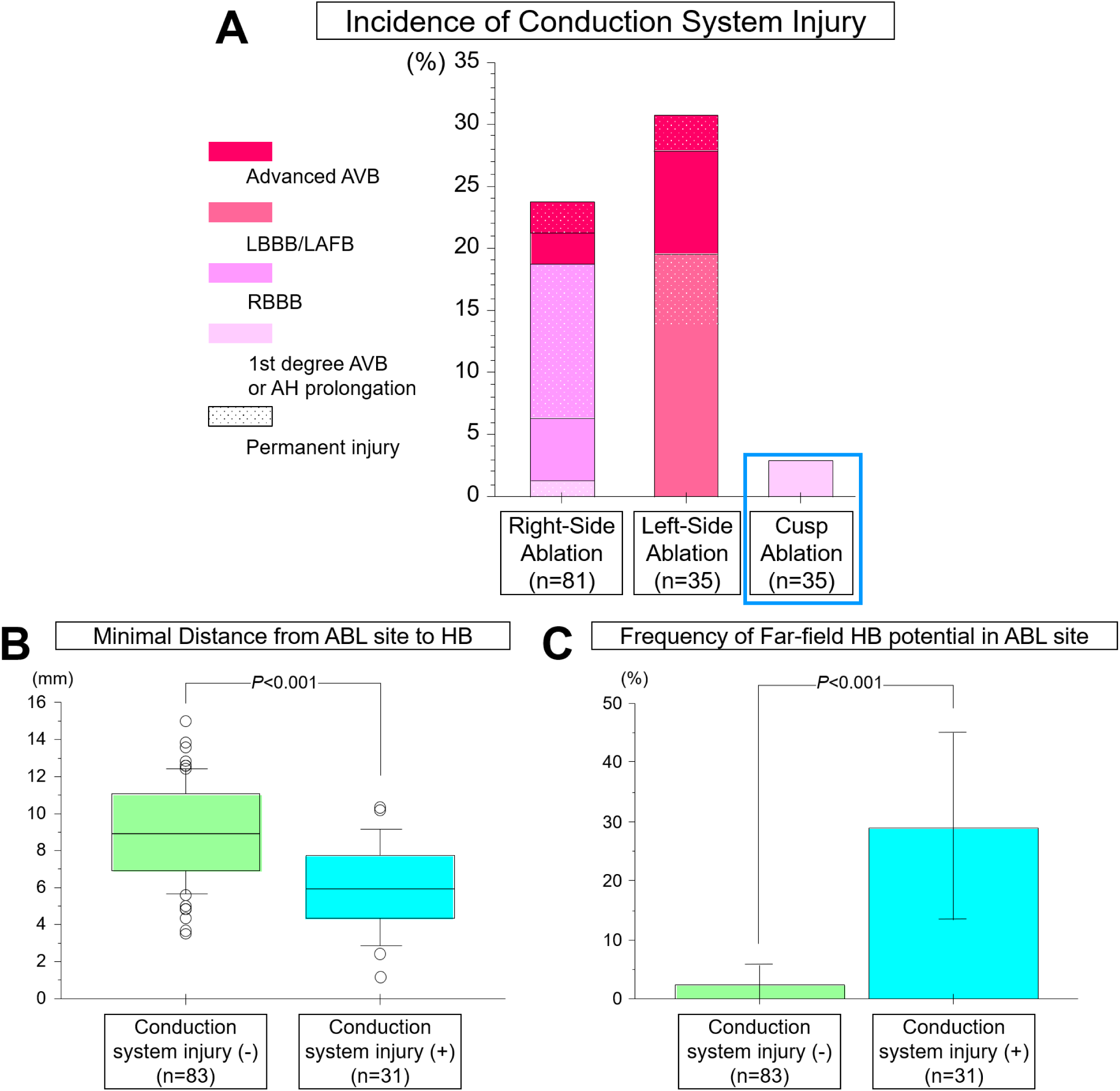
Incidence of conduction system injury during ablation for para-Hisian PVC. **(A)** Frequency of conduction system injury among three ablation sites. Only one case of AH prolongation was observed among a total of 35 cusp ablations (Blue box). **(B)** Comparison of the minimal distance from the ablation site to the nearest His bundle (HB) potential recording site. **(C)** Comparison of the frequency of far-field HB potential detection at the ablation site. AVB indicates atrioventricular block; LAFB, left anterior fascicular block; LCC, left coronary cusp; RBBB, right bundle branch block; and other abbreviations as in **Figure 1**.

In the 31 cases with conduction system injury, the minimal distance from the ablation site to the closest HB potential recording site was significantly shorter than in those without injury (5.9 ± 2.6 mm vs. 8.9 ± 2.7 mm; *P* < 0.001) (**Figure 7B**). Far-field HB potential was also more frequently recognized at the ablation site when conduction injury occurred (29.0% [9 of 31] vs. 2.4% [2 of 83]; *P* < 0.001) (**Figure 7C**).

Figure 8 illustrates the only case in which conduction system injury occurred during cusp ablation. Anatomically, the RCC was in close proximity to the HB, with a minimal distance of 4.9 mm. A far-field HB potential was recorded on the distal bipolar EGM at the ablation site at the RCC. Although PVCs were successfully terminated by RF ablation at the RCC, the AH interval progressively prolonged from 99 to 142 ms, with subsequent recovery to baseline by the end of the procedure.

**Figure 8.**
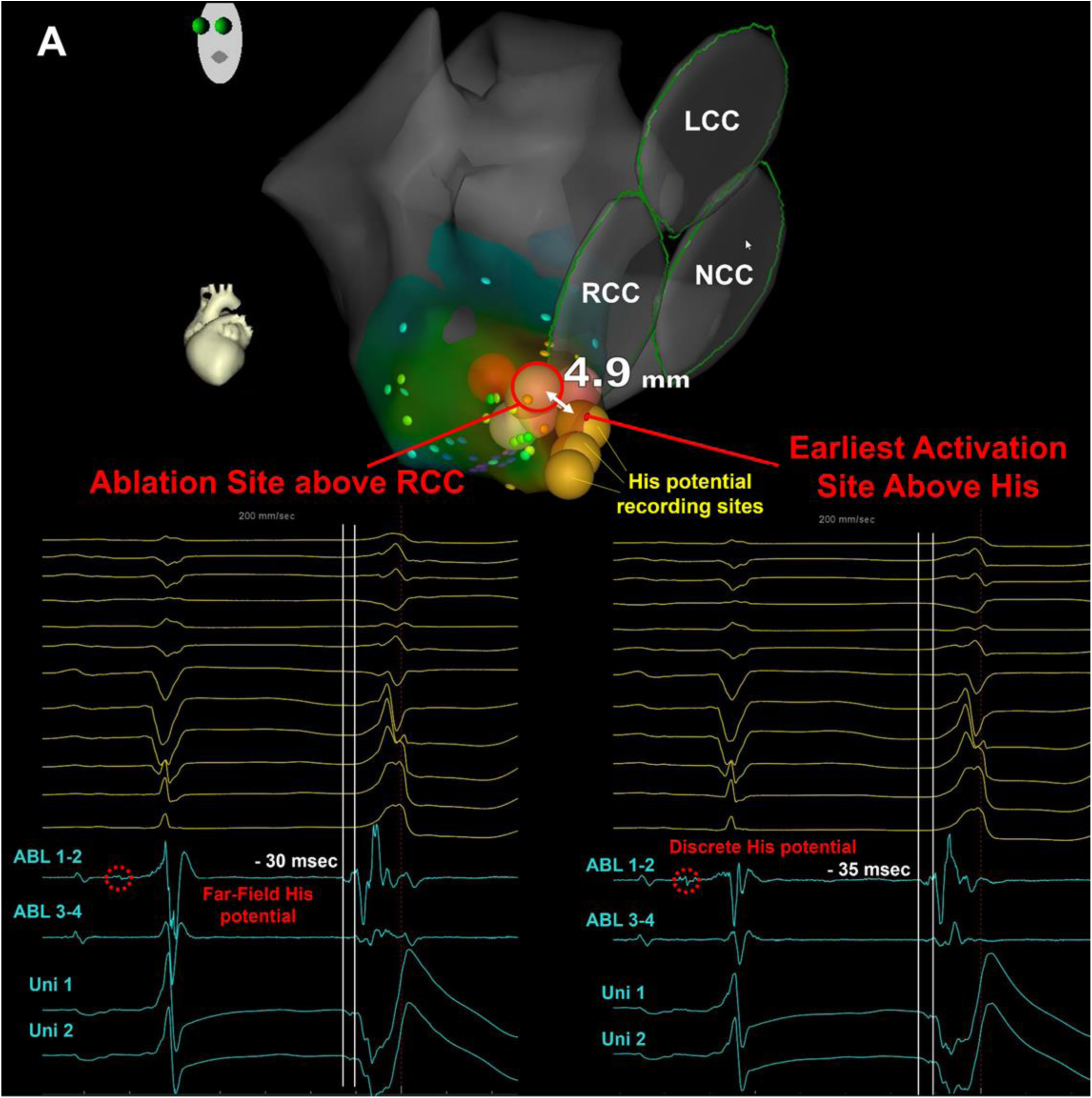
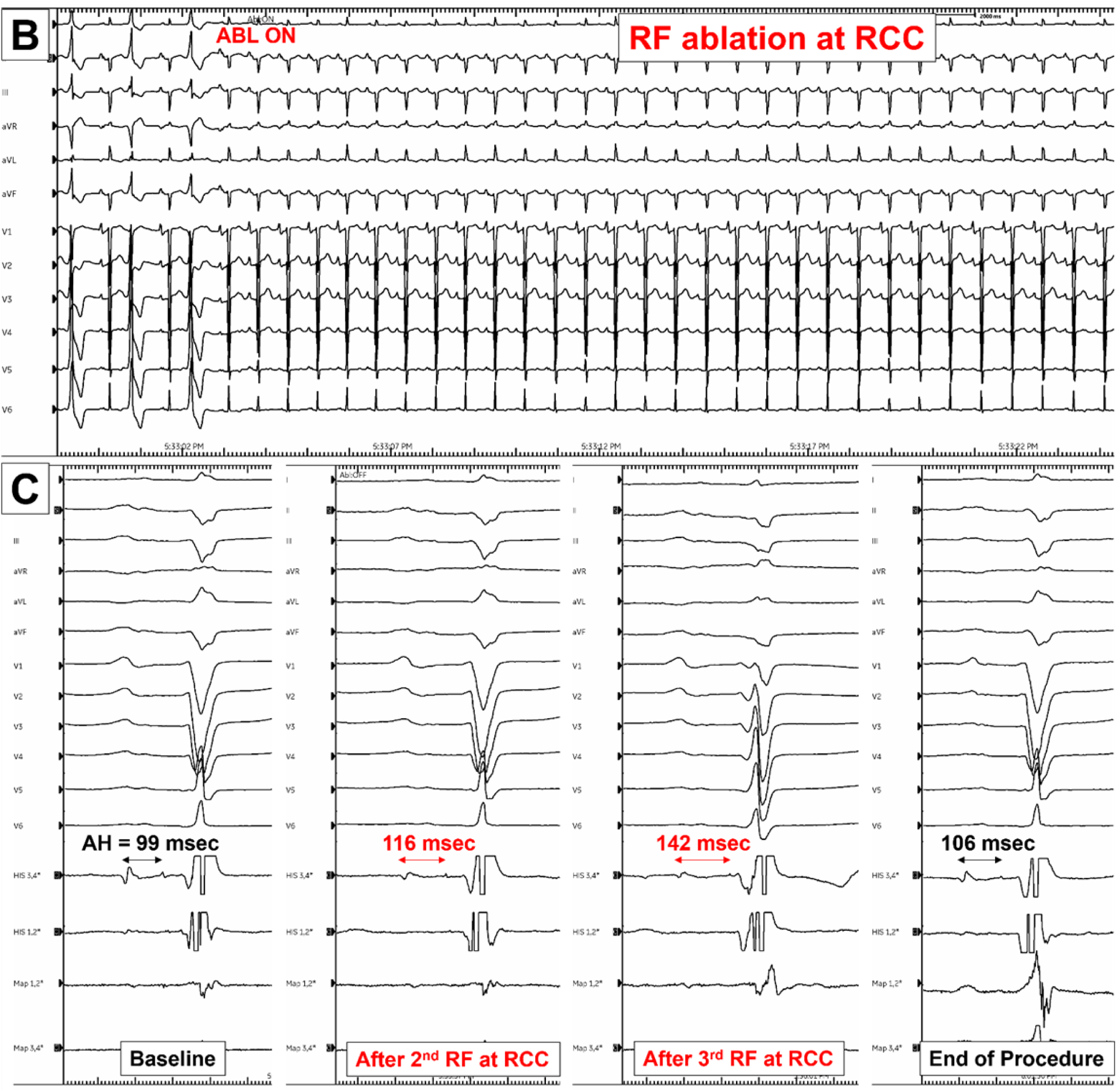
A representative para-Hisian PVC case in which cusp ablation caused transient AH prolongation. See the main text for the details. LCC indicates left coronary cusp; RF, radiofrequency; and other abbreviations as in Figure 1.

## Discussion

To the best of our knowledge, this represents one of the largest retrospective series of ablations for para-Hisian PVC, with a particular focus on the anatomical ablation approach from the aortic coronary cusps, evaluating the procedural success and complications. The main findings in this study are as follows:

1. Catheter ablation of para-Hisian PVCs achieved an acute procedural success rate of approximately 70%, however, nearly half of these cases experienced PVC recurrence during follow-up.
2. Patients with cusp ablation demonstrated a better outcome at the follow-up phase compared with patients without cusp ablation (85% vs. 42%, *P* = 0.005, Figure 4). Ablation at the coronary cusps affected PVC by eliminating, temporarily suppressing, or changing morphology in 80% of cases, suggesting a contribution of cusp ablation to PVC suppression.
3. Earlier activation timing at the cusp and shorter anatomical distance between the cusp ablation site and the EAS in the HB region were predictive of successful cusp ablation. Additionally, the anatomical location of the EAS relative to the HB (above the HB) predicted a procedure success by the anatomical approach.
4. Conduction system injury, including temporary and permanent, occurred in approximately 30% of cases involving HB-region ablation, while only one transient AH interval prolongation was observed during cusp ablation, highlighting the safety of cusp ablation.

In this study, the acute procedural success rate of ablation for para-Hisian PVCs was <80%, and even among patients who achieved acute success, approximately half failed to maintain adequate PVC suppression during follow-up. Also, conduction system injury occurred in approximately 30% of cases. These findings highlight that ablation of para-Hisian PVCs remains highly challenging. In addition to their close proximity to the cardiac conduction system, para-Hisian PVCs frequently originate from intramural sites within the mid-myocardium. ^5,12^ These factors may contribute to procedural complexity. Given the intramural origin, previous studies have emphasized the need for ablation from multiple anatomical sites in many para-Hisian PVC cases.^3,4,19^

In this present study, mappings and ablations were performed from multiple anatomical locations, including right-side HB, left-side HB, and coronary cusps, as described in detail in **Supplemental** Figures 1–4. Although approximately half of the patients experienced recurrence during follow-up (Figure 2D, **Table 1**), patients with cusp ablation had a significantly higher success rate compared with patients without cusp ablation at follow-up (85% vs. 42%, *P* = 0.005, Figure 4**, Supplemental Figures 3 and 4**). This suggested that an anatomical approach from the coronary cusp may improve long-term procedural outcomes.

### Efficacy of ablation from the coronary cusps for para-Hisian PVC

The posterior part of the RCC is adjacent to the central fibrous body, which carries the penetrating portion of the HB. Anteriorly, the RCC is related to the bifurcating AV bundle and the origin of the left bundle branch. The NCC lies superior to the central fibrous body. The HB penetrates through the central fibrous body and continues as the AV conduction bundle that then passes to the crest of the muscular ventricular septum, immediately beneath the membranous septum. Given this close anatomical relationship, Yamada et al. first reported that ablation from the aortic cusp can be effective in a subset of para-Hisian PVCs,^8^ which has since been supported by several subsequent studies. ^5–9^

In the present study, successful ablation from the aortic cusp has been observed in cases exhibiting distinct electrophysiological and anatomical characteristics, including (1) early activation in the cusp region, (2) an “above-HB” earliest activation site, and (3) a short distance between the EAS in the HB region and the cusp (Figures 5B **and 5C**). These features should be carefully assessed when considering a cusp-based ablation approach. Even when the activation timing at the aortic cusp is comparable to that at the EAS, ablating one of several potential exit sites may still enhance overall procedural efficacy, as illustrated in the representative case (Figure 6). This observation aligns with the findings reported by Enriquez et al., supporting the concept that targeted ablation at anatomically adjacent exit sites can contribute to improved outcomes in para-Hisian PVCs. ^3^

### Incidence of conduction system injuries: direct ablation vs. coronary cusp ablation

Conduction system injury, including minor forms, was observed in approximately 30% of cases when RF energy was applied near the HB area (Figure 7A). Both the anatomical proximity to the HB and the presence of far-field HB recorded at the ablation site were associated with an increased risk of conduction injury (Figures 7B **and 7C**). Notably, conduction system injuries were also observed in cases with adequate distance (e.g., 8–10 mm) without far-field potentials in this study. Para-Hisian PVCs are typically defined as those originating within 10 mm of an HB potential recording site, underscoring that ablation within this region inherently carries a substantial risk of AV conduction disturbance.

On the other hand, as observed in this study, ablation from the aortic cusp was associated with a markedly lower incidence of conduction system injury compared with direct ablation at the HB region (Figure 7A). The lower incidence of conduction system injury observed during coronary cusp ablation may be explained by several factors: (1) improved catheter stability within the coronary cusps during RF energy delivery; (2) the anatomical separation between the conduction system, which lies inferior to the coronary cusp leaflets, allowing energy to be delivered indirectly to the PVC origin through the aortic cusp leaflet; and (3) the ability to more readily uptitrate energy delivery while carefully monitoring for conduction system injury, facilitated by the stable catheter position. As demonstrated in one of the present cases (Figure 8), only a single patient exhibited transient AH interval prolongation, which was promptly recognized, allowing for the immediate termination of energy delivery due to the stable catheter position with a clear EGM on the ablation catheter.

### Advantages of mapping and ablation in the coronary cusps: clinical implications

Given the potential intramural origin of para-Hisian PVCs, ablation from multiple anatomical locations is often required to enclose the exit sites. ^3,4^ In this context, the observation that coronary cusp ablation affected PVC resulting in transient PVC suppression or a change in PVC morphology (not only eliminating PVC) in 80% of patients is a clinically important finding. As actually demonstrated in this study, the outcome of ablation was significantly improved when coronary cusp ablation was incorporated. This was further pronounced when the EAS in the HB region was located close to the cusps (above the HB), as shown in Figure 5B **and 5C**. Moreover, even in cases in which the local EGM at the HB region was equivalent to or even slightly later than the earliest activation site, ablation could still be effective or result in a change in PVC morphology, as illustrated in Figure 6 (the cusp area was slightly later than the right-side HB area in this case). Importantly, ablation from the coronary cusps offers a safer alternative to direct ablation near the HB region, with a lower risk of conduction system injury as shown in this study.

Taken together, these findings support the consideration of mapping and ablation from the coronary cusps as an integral component of the ablation strategy for para-Hisian PVCs, particularly when the EAS is located above the HB region.

### Limitation

Several limitations should be acknowledged. First, this was a retrospective study conducted over a 10-year period, during which mapping and ablation strategies were not standardized across cases. In addition, a non-negligible proportion of patients was lost to follow-up. A prospective, randomized controlled trial would be necessary to establish the optimal ablation approach for para-Hisian PVCs. Second, procedural outcomes may have been influenced by factors such as catheter contact, energy delivery, and power settings; however, these parameters could not be fully controlled or uniformly compared in this analysis. Third, the spatial resolution of three-dimensional EAM depends on both the mapping catheter characteristics and overall point density, which may affect the precise localization of the EAS and the HB.

## Conclusions

Although ablation of para-Hisian PVCs remains technically challenging, ablation from the aortic coronary cusps may represent a viable alternative, offering a favorable safety profile and potentially improved efficacy. This approach should be considered after careful assessment of PVC activation timing and the anatomical relationship between the aortic cusps and the HB region.

## Data Availability

The data that support the findings of this study are available from the corresponding author upon reasonable request.

## Acknowledgment

The authors thank Bryan Mitchell (Johnson&Johnson MedTech), Matt Baumann (Abbott) and Monica Korzon (Abbott) for their kind support in analyzing the 3D mapping.

## Nonstandard Abbreviations and Acronyms

AVB: atrioventricular block
EAM: electro-anatomical mapping
EAS: earliest activation site
EGM: electrogram
HB: His bundle
LAFB: left anterior fascicular block
LBBB: left bundle branch block
LCC: left coronary cusp
NCC: non-coronary cusp
PVC: premature ventricular contraction
RBBB: right bundle branch block
RCC: right coronary cusp
RF: radiofrequency

